# Bayesian Model Averaging to Account for Model Uncertainty in Estimates of a Vaccine’s Effectiveness

**DOI:** 10.1101/2021.05.12.21257126

**Authors:** Carlos R Oliveira, Eugene D Shapiro, Daniel M Weinberger

**Affiliations:** Department of Pediatrics, Section of Infectious Diseases and Global Health, Yale School of Medicine, P.O. Box 208000, New Haven, CT, 06520-8000, USA; Department of Biostatistics, Section of Health Informatics, Yale School of Public Health, P.O. Box 208034, New Haven, CT, 06520-8034, USA; Department of Epidemiology of Microbial Diseases, Yale School of Public Health, P.O. Box 208034, New Haven, CT, 06520-8034, USA

**Keywords:** Bayesian Model Averaging, Model Uncertainty, Vaccine Effectiveness, Lyme Vaccine

## Abstract

Vaccine effectiveness (VE) studies are often conducted after the introduction of new vaccines to ensure they provide protection in real-world settings. Although susceptible to confounding, the test-negative case-control study design is the most efficient method to assess VE post-licensure. Control of confounding is often needed during the analyses, which is most efficiently done through multivariable modeling. When a large number of potential confounders are being considered, it can be challenging to know which variables need to be included in the final model. This paper highlights the importance of considering model uncertainty by re-analyzing a Lyme VE study using several confounder selection methods. We propose an intuitive Bayesian Model Averaging (BMA) framework for this task and compare the performance of BMA to that of traditional single-best-model-selection methods. We demonstrate how BMA can be advantageous in situations when there is uncertainty about model selection by systematically considering alternative models and increasing transparency.

## INTRODUCTION

Observational studies are the most appropriate way to measure a vaccine’s effectiveness (VE) post-licensure [1]. When the vaccine-preventable disease is rare, or when there is a prolonged latency between immunization and disease, the case-control design can be particularly favorable, as a smaller sample size is needed and subjects do not need to be followed for a prolonged period of time [2]. As an observational method, however, the case-control design is susceptible to the effects of confounding [3]. The ideal time to deal with known confounders is during the design phase of the study. However, assessment for and control of confounding is often also needed during the analyses of the study, which is most efficiently done using multivariate modeling [4].

Multivariable modeling’s primary goal is to determine whether the observed effect of vaccination on disease is confounded by one of the measured variables, and thus, requires some form of adjustment. The simplest approach assumes that all collected variables are important and uses the “full model,” which includes all potential confounders as independent variables. However, the statistical efficiency of the study will be significantly reduced when unimportant or non-confounding variables are included in a model [5]. More importantly, certain variable combinations may introduce bias due to collider stratification [6], which can lead to erroneous conclusions regarding statistical significance [7, 8].

To address the problems of over-adjusting, several strategies have been developed that help investigators select the “best” model among a set of candidate models and estimate the effectiveness with the assumption that the chosen model is the best for their data [9]. While these methods are easy to implement, they can have several limitations. For example, restricting the analysis to a single model can lack transparency, and may lead to uncertainty about the potential effects of the variables ultimately not included in the final model. An alternative approach to selecting a single model is Bayesian Model Averaging (BMA). Several studies have shown that in the presence of model uncertainty, BMA can provide better inferences than single-model selection methods [10, 11]. However, few have explored the use of BMA in the context of case-control VE studies [12, 13]. In this study, we explore the importance of model uncertainty by re-analyzing data from a Lyme VE study using various confounder selection techniques. We then describe an intuitive BMA framework that is tailored for the analysis of case-control VE studies and compare the performance of this BMA approach to that of traditional single-best-model-selection methods.

## MATERIALS AND METHODS

### Vaccine effectiveness data

A vaccine for Lyme disease (LYMErix™) was available for adults from 1998 until 2002. To explore the importance of modeling uncertainty in VE studies, we re-analyzed data from a matched case-control study that aimed to assess the effectiveness of the Lyme vaccine post-licensure. The methods for the Lyme VE study have been previously described [14]. Briefly, cases were residents of Connecticut, 15-70 years of age, reported to the Department of Public Health as having Lyme disease. Up to 2 controls were identified and matched to each case by age. All participants were interviewed, and medical records were reviewed from primary care providers to ascertain immunization history, evaluate potential confounders, and verify the participant’s case-control status. Study definitions are summarized in Supplementary Table S1.

### Statistical analysis

Unadjusted and adjusted odds ratios (OR), along with their 95%CI, were estimated using conditional logistic regression. The VE was calculated as (1 − OR) x 100%. We first considered the unadjusted model, which contained only the dichotomous immunization status variable, and the full model, which adjusted for all potential confounders. We then tested 3 traditional, single-model-selection approaches. The first selection approach was the two-stage method. In this approach, we first considered each of the potential confounders separately by fitting numerous binary conditional logistic regression models. A multivariable model was then built by including all variables that were statistically significant (p-value <0.05) on the binary models. The second selection approach we used employs backward selection. This approach began with the “full model” (all potential confounders), and then each potential confounder was eliminated, one-at-a-time (least significant first), based on whether it had a p-value cutoff of <0.2 [15]. The third approach we used incorporated the leaps-and-bounds algorithm to search across the modeling space and select the model with the lowest Akaike information criterion (AIC) score [16].

### Bayesian Model Averaging Framework

In the Bayesian framework, it is asserted that there is uncertainty about what the best model is. Hence, our approach begins by first assigning a prior probability to each model to create a probability distribution of all candidate models. The prior probabilities reflect how likely a given model is based on how well it fits the data and how many covariates exist in each model. To approximate this, we use the Bayesian information criterion (BIC), which is a measure of goodness of fit that penalizes the overfitting models (based on the number of parameters in the model), and minimizes the risk of multicollinearity [17, 18]. BIC estimates can be obtained in most statistical packages or can easily be computed with two values from the logistic regression outputs (sample size and the maximum likelihood).

Our approach entails systematically searching for potential models using every combination without repetition of non-collinear control variables. After fitting each potential model, we assign a BIC score to each model and approximated the probability of each model by comparing its fit and complexity relative to the “best BIC-model” (the model with the lowest BIC). Model weights were then estimated using the relative difference in BIC’s, which is equivalent to the Bayesian posterior model probability given the candidate models. The BIC-derived weights are then used to rank all models by their posterior probability of being the best approximation of the true model. We use this rank to ascertain how informative each candidate model is relative to the top model. Then, we use the estimated posterior model probabilities to reduce the modeling space to the most realistic models by dropping the models that provide little to no improvement in fit (i.e., those with a posterior probability of <1% relative to the best model). After reducing the modeling space to only the top subset of models, we recompute the BIC-weights for the newly defined subset of models and only use this top subset for model averaging procedures. Credibility intervals (95%CI) are calculated using the unconditional error. After calculating the pooled OR and 95%CI on the log scale, they are exponentiated to obtain the weighted average of the VE estimate. This BMA approach also allows for comparisons to be made about the relative importance of each potential confounder based on how it may improve the fit of the model. To make these assessments, we extract the BIC-weights from the top subset of candidate models that included a confounder of interest and take the sum of them to estimate the given variable’s posterior inclusion probability (PIP). For additional details see Supplemental Methods.

## RESULTS

### Lyme vaccine effectiveness

A total of 985 subjects were included in the analysis (397 cases and 588 matched controls) of the parent Lyme VE study. Data from a total of 13 potential confounders were considered for inclusion in the multivariate models as potential covariates. The distribution of potential confounders by case status is shown in Table 1. An initial set of 8,192 distinct candidate models were fit and considered for the analyses, ranging from the simplest model (only the vaccine variable) to the most complex model (the vaccine variable plus all potential confounders). The un-weighted distribution of the VE across all models ranged between 56% and 73% (Supplementary Figure S1).

**Table 1.** Characteristics of Participants Enrolled in Lyme Vaccine Study by Case Status, N=985.

| **Variables** | **Cases, N (%)** |  | **Matched Controls, N (%)** |  | **P value <sup>1</sup>** |
| --- | --- | --- | --- | --- | --- |
| Total | 397 | (40%) | 588 | (60%) | - |
| Demographics |  |  |  |  |  |
| Age (yrs.), median (range) | 46 | (15-70) | 47 | (13-69) | 0.88 |
| Non-Hispanic White | 386 | (97%) | 574 | (98%) | 0.35 |
| Female | 208 | (52%) | 394 | (67%) | <0.01 |
| Some college education | 278 | (70%) | 452 | (77%) | <0.01 |
| Currently unemployed | 46 | (12%) | 75 | (13%) | 0.41 |
| Risk Factors |  |  |  |  |  |
| Has an occupational exposure to ticks | 58 | (15%) | 55 | (9%) | <0.01 |
| Lives near wooded area | 375 | (94%) | 544 | (93%) | 0.21 |
| Engages in frequent outdoor activities | 337 | (85%) | 479 | (81%) | 0.12 |
| Has pets at home | 247 | (62%) | 391 | (66%) | 0.02 |
| Protective Measures |  |  |  |  |  |
| Uses protective clothing while outdoors | 177 | (45%) | 346 | (59%) | <0.01 |
| Use of tick repellent on skin or clothing | 117 | (29%) | 196 | (33%) | 0.03 |
| Performs checks for ticks post-exposure | 296 | (75%) | 443 | (75%) | 0.82 |
| Sprays property with tick acaricides | 14 | (4%) | 40 | (7%) | 0.01 |

<sup>1</sup> p-value calculated using bivariable conditional logistic regression.

### Bayesian model averaging

After exclusion of the non-informative models (those with a probability of being the best model <0.01), the top subset of candidate models was selected (n=15), and weights for each model in the top subset were re-normalized for model averaging procedures. The VE estimates for the top 15 models and the consensus BMA estimates are shown in Figure 1. For comparison, the estimate using a model that only controlled for race, which was excluded from model averaging procedures due to its poor fit, is also shown in Figure 1. Averaging across the top 15 models, the model-averaged aOR was 0.31, corresponding to a VE of 69% (95%CI= 18% to 88%). The components of each model, their BIC-derived weights (i.e., their model probability), and the PIP of each potential confounder are summarized in Figure 2.

**Figure 2.**
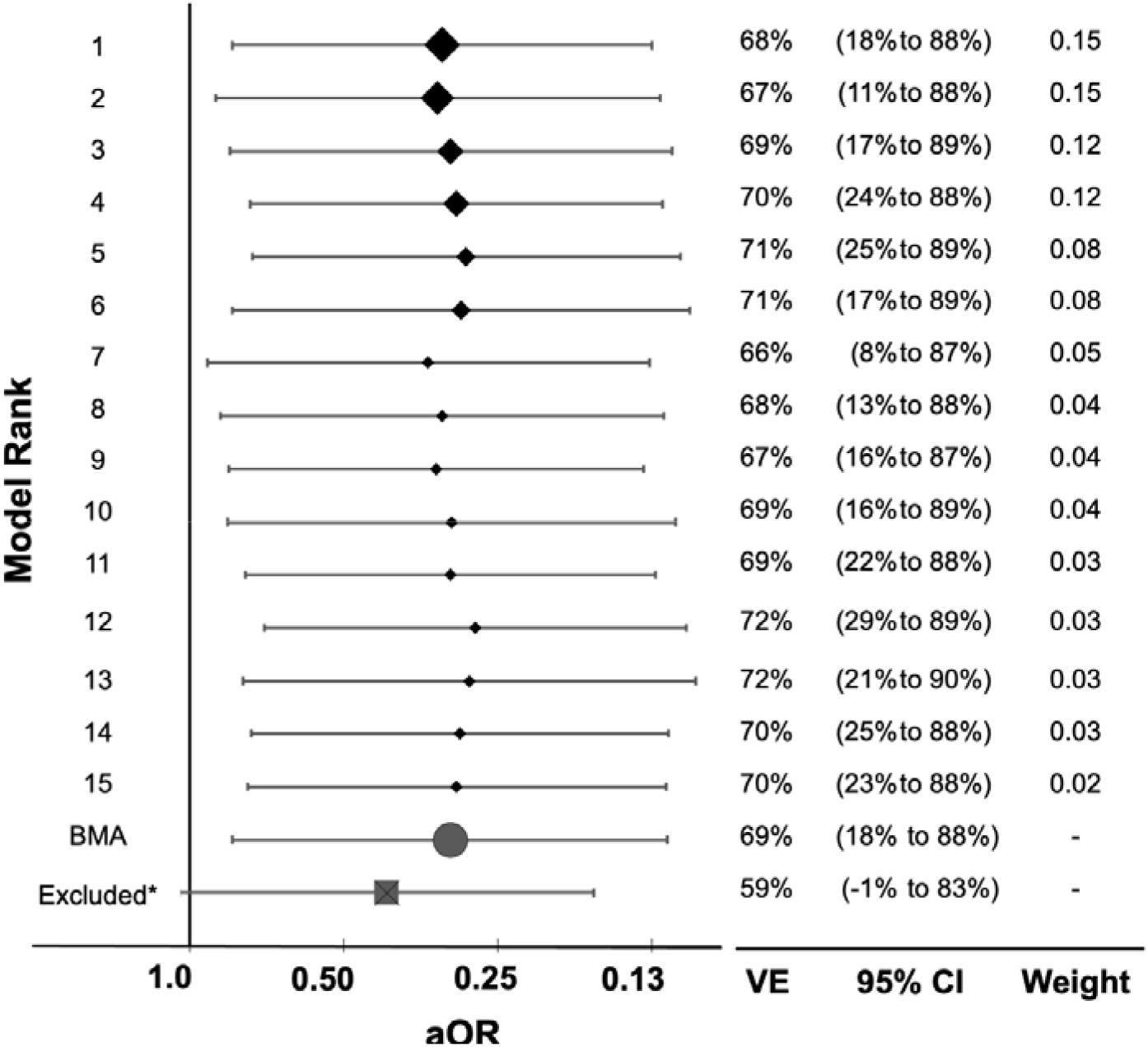
Vaccine effectiveness for model-averaged and the top subset of candidate models. *Excluded from BMA if PIP<0.01; aOR= adjusted odds ratio; VE = vaccine effectiveness; BMA = Bayesian model averaging; PIP = posterior inclusion probability

### Comparison of model selection approaches

Figure 3 summarizes the performance of the various single-best model-selection techniques examined. The unadjusted effectiveness of 3 doses of Lyme vaccine was 59% (95% CI: -4% to 84%). On bivariate analysis, we identified seven variables that were significantly different between cases and matched controls based on P-values. Controlling for those confounders in a multivariable analysis (i.e., two-stage method), the estimated adjusted effectiveness of 3 doses of the vaccine was 71% (95% CI: 21-90%). The estimate of VE using the backward stepwise elimination approach was 73% (95% CI: 26-90%), slightly higher than the two-stage approach. The estimates of VE using the leaps and bounds algorithm and selecting the best model based on AIC was 74% (95% CI: 27-91%), which most closely resembled the estimate of the full model (VE=75%; 95% CI: 28-91%). Between all of the confounder selection approaches tested, the BMA approach resulted in the lowest standard error (SE=0.499).

**Figure 3.**
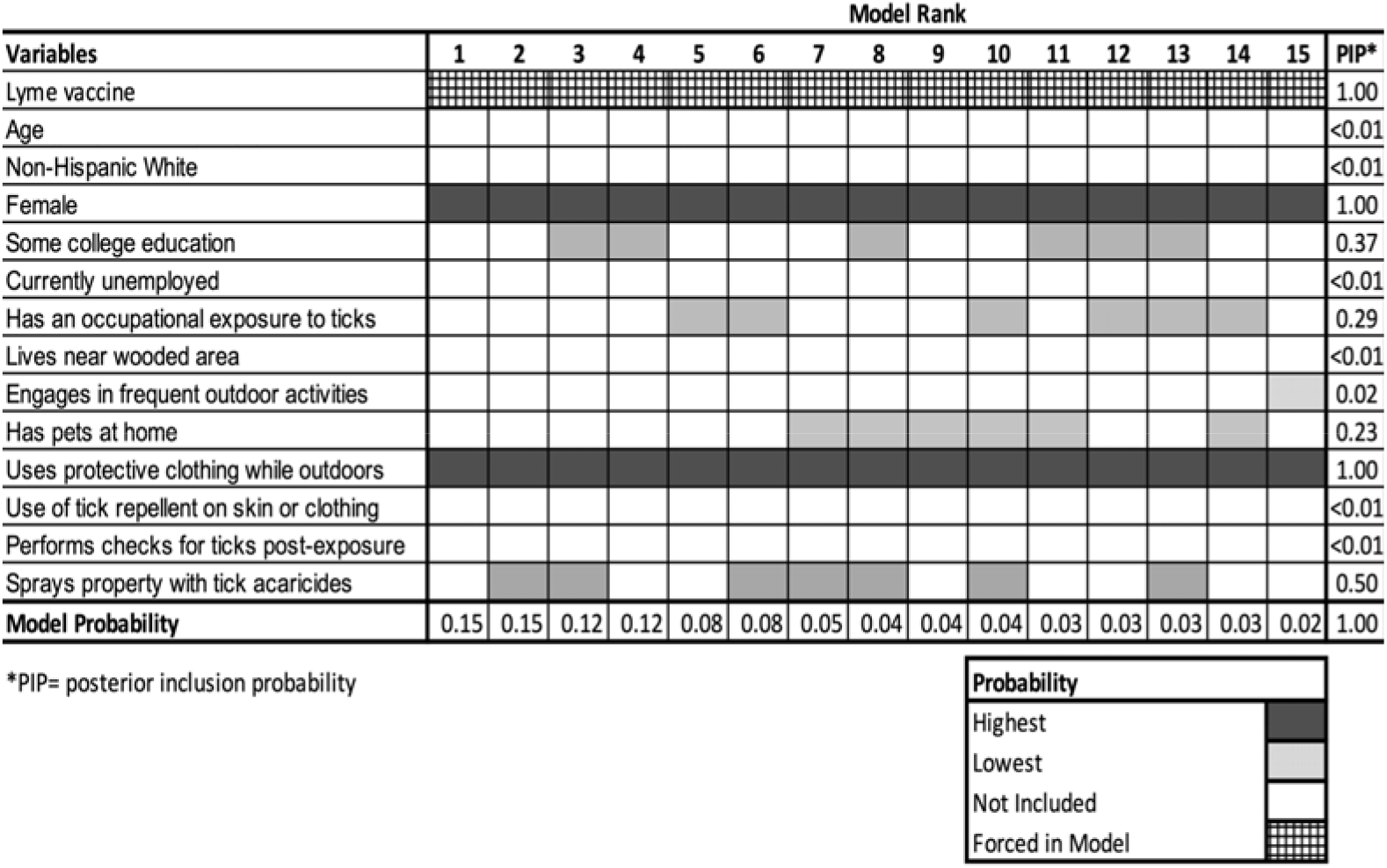
Composition and posterior inclusion probabilities of confounders among the top subset of models. In this heat map, each row corresponds to a potential confounder, and each column corresponds to a particular model. The best model is on the left, and the worst model is on the right. The shade of each square represents the posterior probability of each potential confounder. The darker shades represent higher posterior probabilities, and unshaded squares represent variables that were not included in a given model. Since the immunization status was included in all models a priori, it is checkered, and the PIP for this variable is set to 1.

**Figure 4.**
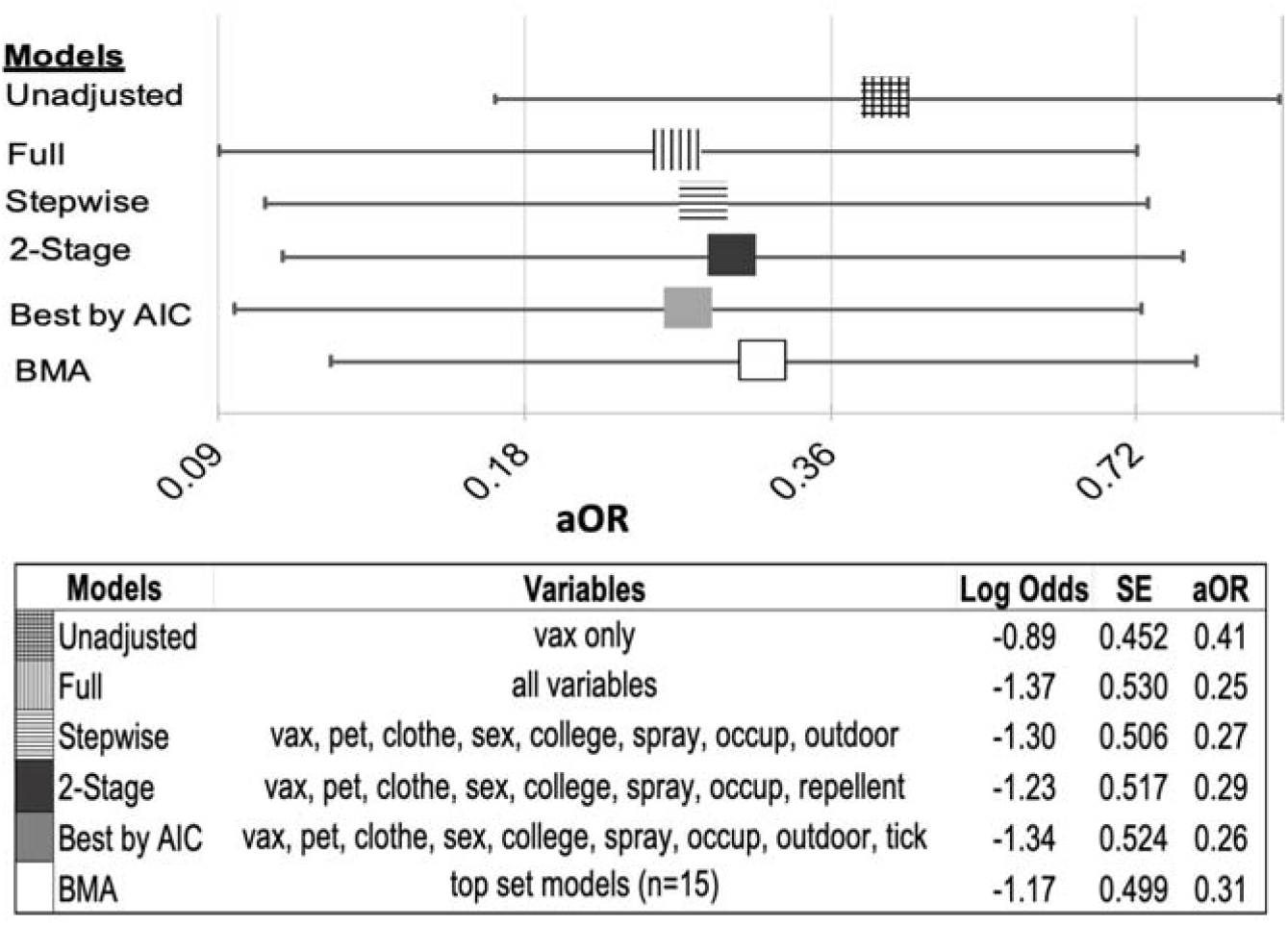
Comparison of single-model selection and Bayesian model-averaging approaches. Vax = vaccine status; Pet = has pets at home; Clothe = wears protective clothing while outdoors; Sex = male or female; College = at least some college education; Spray = sprays property with acaricide; Occup = has an occupational exposure to ticks; Outdoor = engages in frequent outdoor activities; Repellent = use of tick repellent on skin or clothing; Tick = use of tick repellent on skin or clothing; SE = standard error; aOR= adjusted odds ratio; BMA = Bayesian model averaging; AIC = Akaike information criterion

## DISCUSSION

Though case-control VE studies have been conducted for decades, little prior work has been done to define the optimal approaches for variable selection in this context. In this article, we explore the value of using BMA in the analysis of case-control VE data. We show how in addition to enhancing transparency and accounting for model-selection uncertainty, BMA procedures tend to outperform traditional single-best model selection methods in terms of standard errors.

Several additional key insights can be gained from this study. First, we show how the selection of a poorly specified model could significantly affect the interpretation of the VE. For example, when re-analyzing the Lyme VE data, had the model that only controlled for race been selected as the final model (excluded model in Figure 1), the VE would have been significantly lower (59%), and the confidence intervals would have been bloated to suggest that it was not statistically significant (95% CI: -1% to 83%). This demonstrates the potential advantage of BMA, as it is systematic in the consideration of alternative models and can facilitate assessments of the effect confounders have when included in the model [19].

Second, we show how in the context of uncertainty, BMA methods are likely to provide a more complete picture than single-model-selection methods. We found that the top-ranked model in the Lyme VE study had a weight of 0.15, which means that there is a 15% probability that it is the best model given the alternative models. Had this model clearly stood apart from all candidate models as the one with the best fit (i.e., if it is >90% better than all other candidate models), then further BMA would have been unnecessary, and the single best model could have been used to estimate the VE. However, the models ranked 2-4 had a similar fit and competed for the top spot (probabilities of 0.12-0.15). This variability underlines another advantage of using the BMA estimate, as this uncertainty in model selection is then accounted for in the credibility intervals across several models [20].

In this paper, we show how a BMA analytic approach can provide greater transparency in publication by offering an avenue for the reporting of the VE’s distribution under numerous hypothetical scenarios. The BMA procedures outlined in this study are intuitive and, by using BIC-derived weights, are easy to implement with most statistical software. Though this framework does not rule out the possibility that uncontrolled confounding or systematic biases could have affected the results of the study, by incorporating modeling uncertainty into the parameter estimation and through multi-model inferences, BMA can be used to lend an additional measure of rigor and credibility to a well-designed study.

## Data Availability

Owing to data privacy regulations, the raw Lyme vaccine data for this study cannot be shared.

