## SUPPLEMENTARY MATERIAL for "Bayesian Model Averaging to Account for Model Uncertainty in Estimates of a Vaccine’s Effectiveness"

1. **Supplementary Methods**

*1.1 Analysis of Case-Control Data*

|  | **Cases** | **Controls** |
| --- | --- | --- |
| **Immunized** | a | b |
| **Not immunized** | c | d |
| **Total** | a+c | b+d |

In its most basic form, the odds ratio (OR) from case-control studies can be presented and calculated from a 2 x 2 table

In this form, the unconditional OR is equal to the ratio of the odds of a case being immunized to the odds of a control being immunized (Equation 1).

$$OR= \frac{\mathrm{ad}}{\mathrm{bc}}= \frac{Odds that a case was vaccinated}{Odds that a control was vaccinated}$$

(1)

The VE is calculated as (1 − OR) x 100%. Since no randomization can be done, to control for confounding after the data is collected, investigators must either partition the data into a series of strata or use multivariable statistical modeling. Partitioning or stratifying entails dividing the dataset into a series of subgroups or strata to create homogeneity for each confounding variable. To achieve this, separate 2x2 tables can be constructed to remove the effect of the known confounding variable. If there are multiple confounders that need to be considered, this approach becomes statistically inefficient as each additional stratum decreases the sample size, and subsequently, the statistical power. An alternative approach is to use statistical modeling, particularly logistic regression models, specifically,

$$\text{log }\left( \frac{\text{case}}{\text{control}} \right)\text{=}\beta_{0}\text{ + }\beta_{1}\text{(vax) + }\beta_{2}\text{(cov\_1) + }\beta_{3}\text{(cov\_2) + … + }\beta_{k}\text{(cov\_k)}$$

(2)

where the probability of being a case is a function of the dichotomous variable corresponding to receipt of the vaccine β_1_, and β_2_ through β_k_ are the coefficients for each covariate included in the model to control for confounding. In a matched case-control study, the constant term β_0_ is replaced with an indicator term for being in a specific matched case-control set (i.e., conditional logistic regression).

1.2 *Bayesian Model Averaging Framework*

$$\text{p}\left( \text{M}_{\text{k}} | \text{D} \right)\text{= p}\left( \text{D|M}_{\text{k}} \right)\text{ }\frac{\text{p(}\text{M}_{\text{k}}\text{)}}{\text{p(D)}},$$

(3)

Bayes theorem of total probability describes the probability of observing a particular event based on prior information regarding a related event. This Bayesian reasoning can be extended to the problem of model selection, and the probability that any particular model is the closest approximation to the “true model” (posterior model probability) can be estimated given some prior knowledge about the model. The posterior probability of any given model can then be described as

where $p\left( M_{k} | D \right)$ denotes the probability of model $M_{k}$ given data D, $p\left( M_{k} \right)$ denotes the prior probability of any given model, and $p\left( {D|M}_{k} \right)$ is the integrated likelihood of the model $M_{k}$.

To approximate prior probabilities of each model, we use the Bayesian information criterion (BIC), which can easily be computed from the regression output using the sample size and the maximum likelihood of the model, namely

$$BIC= -2 \times\log\left( \mathrm{likelihood} \right)+\log\left( n \right)\times k$$

(4)

where n is the study sample size, and k is the total number of variables in the given model.

To estimate the posterior probabilities of each model, we divided the BIC of each candidate model over the sum of all the BICs so that they all add up to one. Specifically,

$$\mathrm{wt}_{k}= \frac{\exp\left( \frac{{- \Delta BIC}_{k}}{2} \right)}{\sum_{i=1}^{n} \exp\left( \frac{{- \Delta BIC}_{i}}{2} \right)},$$

(5)

where$\mathrm{wt}_{k}$ denotes the weight for Model k, ${\Delta BIC}_{k}$ is the difference of BIC for Model k and the smallest BIC among all of the candidate models.

The modeling space can be stated mathematically as

$$M_{\mathrm{all}}=\left\{ M_{1}{, M}_{2},\ldots M_{k} \right\}$$

(6)

where M_all_ represents the full modeling space and M_1_ to M_k_ represents each candidate model. How the modeling space should be defined for BMA analyses is subject to much debate. Some early BMA adopters have advocated that model averaging should be performed only on a small number of carefully selected and fully justified model specifications (rather than across all possible models). This is based on the idea that just as uninformative variables decrease the explanatory power of a model, so does inclusion of uninformative models decrease the explanatory power and may even bias the results of the BMA analyses. However, some argue that restricting BMA to only the models that the investigator considers as "most justified" could introduce researcher bias. Setting a model's prior probability to zero a priori may weaken one of the greatest strengths of BMA: the ability to be transparent and systematic in consideration of alternative models. Moreover, systematic and exhaustive searches are more likely than non-exhaustive searches to find the best-fitting models for averaging procedures. Our approach aims to strike a balance between the two views by both conducting an exhaustive systematic search for candidate models yet methodically selecting the best-fitting models for model-averaging procedures. To this end, we first search for potential models using every combination without repetition of non-collinear control variables. Then, we used the estimated posterior model probabilities to reduce the modeling space to the most realistic models by dropping the models that provide little to no improvement in fit (i.e., those with a posterior probability of <1% relative to the best model). Finally, we recompute the BIC-weights for the newly defined subset of models for model averaging procedures. The weighted average of the vaccine coefficient and its standard error is calculated as follows:

| $\hat{V_{x}}= \sum_{i=1}^{2^{k}} {(Vx}_{k})({wt}_{k})$, and | (7) |
| --- | --- |
| $\hat{se}\hat{{(V}_{x}})=\sum_{i=1}^{2^{k}} Var{(Vx}_{k}) \left( {wt}_{k} \right)+\sum_{i=1}^{2^{k}} {({Vx}_{k}-\hat{V_{x}})}^{2} \left( {wt}_{k} \right)$, | (8) |

where${Vx}_{k}$ represents the coefficient for the vaccine variable in Model k, and ${wt}_{k}$ represents the BIC-derived weight of the model. The unconditional standard error combines the uncertainty in both the within-model error $Var{(Vx}_{k})$ and the between-model error $\left( {Vx}_{k}-\hat{V_{x}} \right)^{2}$.

To estimate the posterior inclusion probability (PIP) of each potential confounder, we took the sum of the BIC-weights from all the models that included the confounder of interest. Specifically,

$\Pr\left[ \beta_{1}\neq0 | D \right]= \sum_{M_{k}:Cov\_1\in M_{k}} \left( \mathrm{wt}_{k} \right)$,

(9)

where β_1_, is the coefficient for a potential confounder cov_1. With this approach, if a potential confounder is included in all of the top models, then the PIP will approximate 1 and is equivalent to the probability that it would be included in the top-ranked model.

2. **Supplementary Tables**

**Table S1.** Study Definitions for Study on the Effectiveness of Lyme Vaccine

| Variable | Definition |
| --- | --- |
| Definite Lyme disease | Physician diagnosed erythema migrans measuring > 5 cm, or a positive laboratory test result for antibodies to *Borrelia burgdorferi* and objective evidence of either early disseminated or of late Lyme disease (e.g., facial nerve palsy, meningitis, carditis, or arthritis). |
| Case | Residents of Connecticut, 15-70 years of age, diagnosed with first episode of “definite Lyme disease” between 2000 and 2013. |
| Control | Individuals without signs or symptoms of Lyme disease at focal time, matched to a case by age (±5 years) and geographic area (same telephone exchange). |
| Focal time | The common date between cases and controls. For controls, the focal time will be the date of telephone interview. For cases, the focal time will be the date of their diagnosis of Lyme disease. |
| Exclusion criteria | Significant immunosuppression (because of either an illness or a medication) or prior treatment for Lyme disease. |
| Fully immunized | Received ≥3 doses of Lyme vaccine at least 1 month prior to focal time. |

3. **Supplementary Figures**

**Figure S1.** Distribution of the vaccine’s effectiveness across all possible models**.**

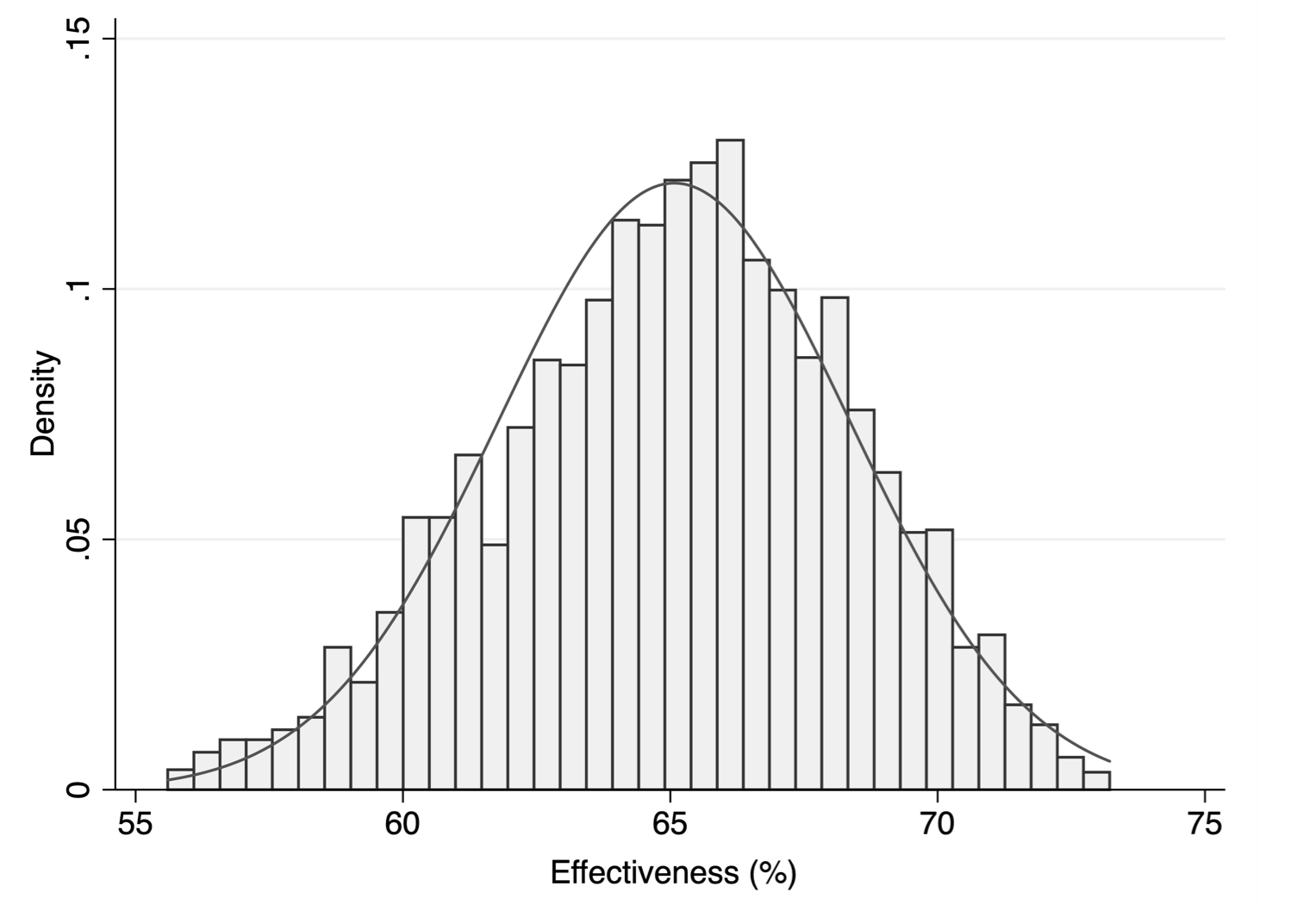
